# Real-time Machine Learning Alerts to Prevent Escalation of Care: A Pragmatic Clinical Trial

**DOI:** 10.1101/2022.12.21.22283778

**Authors:** Matthew A. Levin, Arash Kia, Prem Timsina, Fu-yuan Cheng, Kim-Anh-Nhi Nguyen, Roopa Kohli-Seth, Hung-Mo Lin, Yuxia Ouyang, Robert Freeman, David L. Reich, Sanam Ahmed, Shan Zhao, Rohit Gupta, Madhu Mazumdar, Eyal Klang

## Abstract

**Importance:** Automated machine learning algorithms have been shown to outperform older methods in predicting clinical deterioration requiring escalation of care, but rigorous prospective data on their real-world efficacy are limited.

**Objective:** We hypothesized that real-time deterioration prediction alerts sent directly to front-line providers would reduce escalations.

**Design:** Single-center prospective pragmatic clinical trial conducted from July 2019 to March 2020. The trial was terminated early due to the COVID-19 pandemic. Patients were followed for 30 days post-discharge.

**Setting:** Academic tertiary care medical center located in New York City.

**Participants:** All adult patients admitted to any of four medical-surgical units were included. Assignment to intervention or control arms was determined by initial unit admission.

**Intervention:** Real-time alerts stratified according to predicted likelihood of clinical deterioration sent to the nursing/primary team or directly to the rapid response team. Clinical care and interventions were at the discretion of the providers. For the control units, alerts were generated but not sent.

**Main Outcomes:** The primary outcome was the incidence of escalation of care. Secondary outcomes included orders placed for cardiovascular support, in-hospital and 30-day mortality. Ad-hoc outcomes included time to ICU escalation and time to discharge.

**Results:** 2,780 patients were enrolled, 1,506 in the intervention group and the 1,274 in the control group. Average age was 66.2 years and 1,446 (52%) of participants were female. There was no difference in escalation between the groups, relative risk(RR) 1.22(95% Confidence Interval[CI] (0.97,1.54),p=0.10). Patients in the intervention group were more likely to receive cardiovascular support orders RR 1.35(95% CI (1.10,1.66),p=0.022). Median time to escalation with alerts was 50.6 [21.6-103] versus 58.6 [25.4-115] hours (difference -5.70;95% CI (-10.00,-2.00),p<0.001). The hazard ratio for likelihood of ICU escalation within 12 hours of an alert was 3.36 (95% CI (1.38,8.21),p=0.01) and for faster hospital discharge was 1.10 (95% CI (1.01,1.19),p=0.02). Combined in-hospital and 30-day-mortality was lower in the intervention group, RR 0.72 (95% CI (0.54,0.94),p=0.01).

**Conclusions and Relevance:** Preliminary evidence suggests that real-time machine learning alerts do not reduce the incidence of escalation but are effective in reducing time to escalation, hospital length of stay and mortality.

**Trial Registration:** ClinicalTrials.gov, NCT04026555, https://clinicaltrials.gov

**Key Points:** 

**Question:** Can real-time machine learning generated alerts predicting clinical deterioration, sent directly to front-line providers, reduce escalations in care?

**Findings:** In this pragmatic clinical trial that included 2780 adults, that was terminated early due to the COVID-19 pandemic, the incidence of escalation among patients who received alerts was 11.2% versus 9.7% among patients who did not, a non-significant difference. Combined in-hospital and 30-day mortality in the alerts group was 6.9% versus 9.4% in the group with no alerts, a significant difference.

**Meaning:** Preliminary evidence suggests that real-time machine learning generated alerts do not reduce the incidence of escalation but may reduce mortality.

## Introduction

A key goal of inpatient care is timely intervention to prevent or treat clinical deterioration. Several clinical deterioration prediction scores have been developed, most notably the Modified Early Warning Score (MEWS) and the Royal College of Physicians’ National Early Warning Score.^1–3^ While these scores have been shown to perform well retrospectively, prospective validation has been limited.^4–7^ Recently, automated machine learning approaches trained on large volumes of electronic health record (EHR) data have been shown to outperform older methods.^8–22^ We conducted a pragmatic clinical trial to test the hypothesis that automated real-time alerts, sent directly to front-line providers, would decrease the number of escalations to intermediate care or intensive care units.

## Methods

### Study Setting and Participants

The Realtime Streaming Clinical Use Engine for Medical Escalation (ReSCUE-ME) trial was a single-center prospective pragmatic clinical trial (ClinicalTrials.govNCT number: NCT04026555) conducted at The Mount Sinai Hospital, a tertiary care academic medical center located in New York City. All patients aged 18 or greater admitted to any of four medical-surgical patient units were included in the study. Two of the units received alerts (intervention units) and two did not (control units).

Institutional review board (IRB) approval was obtained (IRB-18-00581) from the Program for the Protection of Human Subjects at the Icahn School of Medicine at Mount Sinai. Written informed consent was waived, based upon minimal risk to participants, since the alerts neither prescribed treatments nor restricted access to the rapid response team (RRT). A description of the study was provided to patients upon admission to the unit and multi-lingual information was posted in prominent locations within each unit. Patients could opt out at any time via a website, telephone call, or direct communication with their provider, nurse, or the unit business associate.

### Machine Learning Model

The predictive model specifics and validation process have been previously described.^23^ Briefly, a random forest model was trained to identify escalation of care or death during an inpatient stay. Features included laboratory test results, vital signs, automated ECG interpretations, and structured clinical nursing documentation (e.g., level of consciousness). Final performance of the retrospective model using the default threshold of 0.5 on a validation dataset of 58,742 patients showed balanced performance with a sensitivity 0.79, specificity 0.79, accuracy of 0.79, and positive predictive value (PPV) of 0.11.

For the prospective trial, silent pilot modeling led to the selection of a threshold of 0.59 for the primary alert and 0.65 for the secondary alert to produce approximately eight primary team and six RRT alerts per day (14 alerts total, seven per intervention unit). The predicted sensitivity was 0.4, specificity 0.78, PPV 0.13, and negative predictive value (NPV) 0.94.

### Study protocol

Primary alerts triggered messaging to the nursing team to perform an assessment and hourly vital signs checks for 4-8 hours. The responsible provider was notified and evaluated the patient to determine whether intervention was warranted. Secondary alerts were sent directly to the RRT to perform a patient evaluation and to provide recommendations to the primary team. In the control units, prediction scores were generated using the same criteria, but no alert notifications were sent; standard clinical care was provided. Both intervention and control units could activate the RRT at any time, irrespective of the alert status. We limited the total number of alerts per patient per unit admission to one to the primary team and three overall to minimize the likelihood of alarm fatigue. Additional alarm fatigue mitigation strategies included suppression of alerts: (1) within two hours following unit admission; (2) if variation from the previous prediction score was less than 10%; and (3) if an alert was sent in the previous eight hours. A detailed process flow diagram of the notification protocol is shown in Supplemental Figure S1.

### Primary and Secondary Study Endpoints

The primary outcome was the incidence of escalation of care from floor to stepdown, telemetry, or ICU. Secondary outcomes per protocol included incidence of orders placed for cardiovascular (CV) support (defined as any order for autonomic, cardiac, cardiovascular or diuretic medications, plus orders for intravenous fluid therapy), combined in-hospital and 30-day mortality, and alert performance as described below. Ad-hoc secondary outcomes included the likelihood of ICU escalation within 12 or 24 hours and the likelihood of earlier hospital discharge.

### Sample size calculation

In a pilot feasibility study, the rate of escalation of care in the selected units was approximately 10%. We expected that real-time alerts would be able to prevent 20% of the predicted escalations (i.e., reduce the rate of escalation to 8%). Using alpha of 0.05 and beta of 0.20, 6,426 subjects, evenly divided between groups, were required to detect such a difference.

### Statistical analysis

Only data from the first hospital admission per patient were analyzed. Multiple admissions to a study unit during a single hospitalization (e.g., returning to floor level care after an ICU stay) were included in the analysis. Continuous variables are presented as mean(standard deviation) or median[interquartile range], as appropriate. Categorical variables are presented as counts(percentage). Student’s two-sample t-test was used to examine the differences in means and Hodges-Lehmann estimation was used to assess differences in median location shift between study arms. The two proportion Z-test was used to examine the differences in proportions between the two study arms. Univariate outcomes were not adjusted for multiple comparisons.

The assignment to intervention or control groups was determined by initial unit admission. Propensity score modeling with stabilized inverse probability of treatment weight (IPTW) was used to account for differences in the baseline characteristics of patients between groups.^24^ The propensity score model predictors were age, sex, race, ethnicity, BMI category, initial prediction score and patient comorbidities as shown in Table 1. An indicator variable for missing comorbidity data was included in the model. Patients with missing comorbidity information (190,6.8%) tended to be younger with lower BMI, and less likely to be escalated or die (Supplemental Table S1).

**Table 1.** Patient characteristics.

|  | **Intervention**<br>**(Received alert)**<br>**(N=1506)** | **Control**<br>**(No alert)**<br>**(N=1274)** | **Difference (95%CI)** |
| --- | --- | --- | --- |
| **Age, years** |  |  |  |
| Mean (SD) | 67.2 (17.0) | 65.2 (17.8) | 2.03 (0.72, 3.33) |
| **Sex** |  |  |  |
| Female | 773 (51.3%) | 673 (52.8%) | -1.5% (-5.3%, 2.3%) |
| Male | 733 (48.7%) | 601 (47.2%) |  |
| **Race** |  |  |  |
| White | 524 (34.8%) | 376 (29.5%) | 5.2% (1.5%, 8.9%) |
| Black | 355 (23.6%) | 267 (21.0%) | 2.5% (-0.9%, 5.8%) |
| Asian | 77 (5.1%) | 95 (7.5%) | -2.6% (-4.6%, -0.6%) |
| Other | 477 (31.7%) | 459 (36.0%) | -5.1% (-8.8%, -1.3%) |
| Missing | 73 (4.8%) | 77 (6.0%) |  |
| **Ethnicity** |  |  |  |
| Hispanic or Latino | 459 (30.5%) | 424 (33.3%) | -3.4% (-7.1%, 0.3%) |
| Not Hispanic or Latino | 974 (64.7%) | 773 (60.7%) |  |
| Missing | 73 (4.8%) | 77 (6.0%) |  |
| **BMI (kg·m<sup>-2</sup>)** |  |  |  |
| Mean (SD) | 27.2 (7.65) | 26.6 (9.10) | 0.58 (-0.07, 1.24) |
| **BMI category** |  |  |  |
| Underweight | 113 (8.0%) | 109 (9.3%) | -1.2% (-3.5%, 1.0%) |
| Normal | 503 (35.6%) | 458 (38.9%) | -3.2% (-7.0%, 0.6%) |
| Overweight | 396 (28.1%) | 321 (27.2%) | 0.8% (-2.7%, 4.4%) |
| Obese | 399 (28.3%) | 290 (24.6%) | 3.7% (0.2%, 7.1%) |
| **First deterioration score on admission** |  |  |  |
| Mean (SD) | 0.56 (0.10) | 0.57 (0.096) | -0.01 (-0.02, -0.01) |
| **Elixhauser score** |  |  |  |
| Mean (SD) | 2.2 (1.4) | 2.2 (1.4) | 0.04 (-0.07, 0.15) |
| **Alcohol use\*** |  |  |  |
| Yes | 72 (5.1%) | 95 (8.1%) | -3.0% (-5.0%, -1.0%) |
| No | 1346 (94.9%) | 1077 (91.9%) |  |
| **Arrhythmia\*** |  |  |  |
| Yes | 60 (4.2%) | 59 (5.0%) | -0.8% (-2.5%, 0.9%) |
| No | 1358 (95.8%) | 1113 (95.0%) |  |
| **Asthma/COPD\*** |  |  |  |
| Yes | 315 (22.2%) | 260 (22.2%) | 0.0% (-3.2%, 3.3%) |

|  | Intervention<br>(Received alert)<br>(N=1506) | Control<br>(No alert)<br>(N=1274) | Difference (95%CI) |
| --- | --- | --- | --- |
| No | 1103 (77.8%) | 912 (77.8%) |  |
| **Dementia\*** |  |  |  |
| Yes | 116 (8.2%) | 106 (9.0%) | -0.9% (-3.1%, 1.4%) |
| No | 1302 (91.8%) | 1066 (91.0%) |  |
| **Depression\*** |  |  |  |
| Yes | 165 (11.6%) | 145 (12.4%) | -0.7% (-3.3%, 1.9%) |
| No | 1253 (88.4%) | 1027 (87.6%) |  |
| **Diabetes\*** |  |  |  |
| Yes | 500 (35.3%) | 432 (36.9%) | -1.6% (-5.4%, 2.2%) |
| No | 918 (64.7%) | 740 (63.1%) |  |
| **Drug abuse\*** |  |  |  |
| Yes | 62 (4.4%) | 59 (5.0%) | -0.7% (-2.4%, 1.1%) |
| No | 1356 (95.6%) | 1113 (95.0%) |  |
| **HIV\*** |  |  |  |
| Yes | 72 (5.1%) | 61 (5.2%) | -0.1% (-1.9%, 1.7%) |
| No | 1346 (94.9%) | 1111 (94.8%) |  |
| **Hypertension\*** |  |  |  |
| Yes | 932 (65.7%) | 735 (62.7%) | 3.0% (-0.8%, 6.8%) |
| No | 486 (34.3%) | 437 (37.3%) |  |
| **Peripheral vascular disease\*** |  |  |  |
| Yes | 134 (9.5%) | 92 (7.8%) | 1.6% (-0.6%, 3.8%) |
| No | 1284 (90.6%) | 1080 (92.2%) |  |
| **Prior malignancy\*** |  |  |  |
| Yes | 423 (29.8%) | 278 (23.7%) | 6.1% (2.6%, 9.6%) |
| No | 995 (70.2%) | 894 (76.3%) |  |
| **Thyroid disease\*** |  |  |  |
| Yes | 208 (14.7%) | 183 (15.6%) | -0.9% (-3.8%, 1.9%) |
| No | 1210 (85.3%) | 989 (84.4%) |  |
| **Missing comorbidity data\*\*** |  |  |  |
| Yes | 88 (5.8%) | 102 (8.0%) | -2.2% (-4.1%, -0.2%) |
| No | 1418 (94.2%) | 1172 (92.0%) |  |

\*Denominator for comorbidity percentages is the number of patients with complete comorbidity data
\*\*Missing comorbidity applies for all comorbidity variables.

IPTW log-binomial regression models were used to model the binary outcomes, including escalations, order placement, and in-hospital and 30-day mortality. Treatment effect is expressed as adjusted relative risk. IPTW Cox proportional hazard regression analysis was used to assess the association of study arm with hospital length of stay. The proportional hazard assumption was validated by testing the study arm by time interaction term in the model (p=0.661). The hazard ratio is used to describe the relative probability of hospital discharge at a given time, with a hazard ratio >1 indicating an association with shorter hospital stay. IPTW Cox proportional hazard regression for clustered analysis was also used to assess whether time to ICU escalation within the first 12 or 24 hours after receiving the RRT alert differed between the two arms. As a patient might have multiple RRT alerts during the hospital stay, each patient was considered a random effect and empirical standard errors of the parameter estimates were used. A hazard ratio >1 indicates an association with shorter time taken to transfer to the ICU. As a confirmatory analysis, we also performed a non-IPTW covariates-adjusted regression analysis. Correction for multiple comparisons was performed on all key outcomes within the secondary hypotheses, and for the ad-hoc analyses, respectively, using the false discovery rate method (FDR).^25^

Alert performance was evaluated by calculating the sensitivity, specificity, PPV, NPV, and accuracy for level of escalation to intermediate care or ICU, and for escalation to ICU only. Secondary analyses were conducted to examine the effects of patients’ presence on the individual units and the relative predictive value of the alert level, due to the bifurcation of order transmission to nursing teams versus RRT. All statistical analyses were performed using R v4.2.1 in RStudio v2022.07.1 (R Foundation, Vienna, Austria). A two-sided p value of <0.05 was considered statistically significant.

## Results

### Patient Characteristics

Following a pilot period of two weeks, the trial began July 2019 and terminated early in March 2020 prior to achieving the targeted sample size due to onset of the COVID-19 pandemic. In total 2,780 patients were enrolled: 1,506 in the intervention group and the 1,274 in the control group (Figure 1). 1,735 alerts were sent to providers for patients in the intervention group, and 1,403 alerts were generated for the control group but not sent. The average total number of alerts sent per day to the intervention units was 6.7: 4.5 primary alerts and 2.2 RRT alerts.

**Figure 1:**
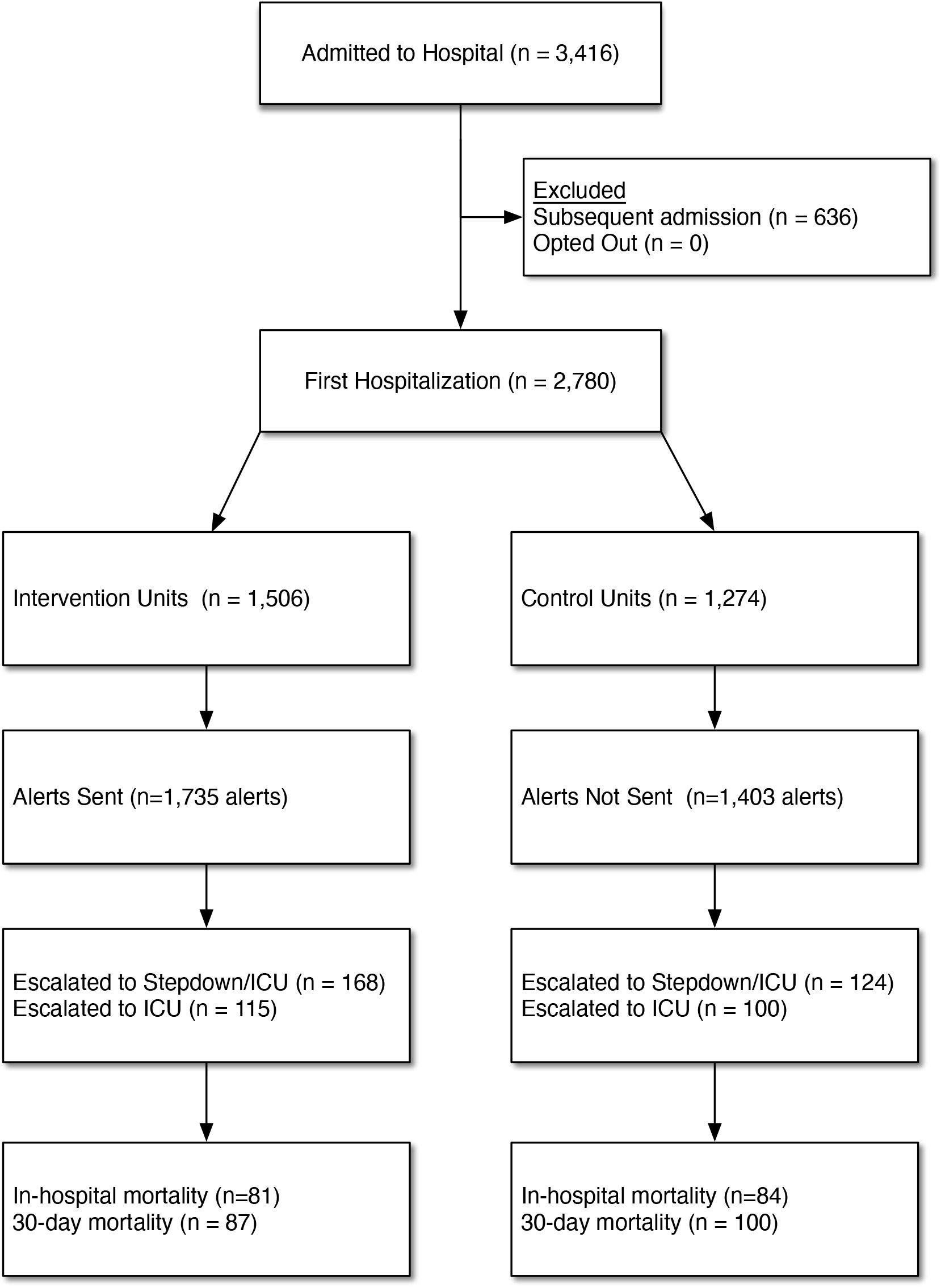
Study Enrollment.

Demographics between groups were similar (Table 1). Clinically relevant differences among patients with complete comorbidity data included higher rates of obesity in the intervention arm, 399/1,506 (28.3%) versus 290/1,274 (24.6%) (absolute difference 3.7%; 95% CI 0.2% to 7.1%), greater history of prior malignancy, 423/1,506 (29.8%) versus 278/1,274 (23.7%) (absolute difference 6.1%; 95% CI 2.6% to 9.6%) and lower rate of alcohol use disorder 72/1,506 (5.1%) versus 95/1,274 (8.1%) (absolute difference -3.0%; 95% CI -5.0% to -1.0%). The patients enrolled were diverse: 22.4% Black, 6.2% Asian, 32.4% White, and 31.8% reporting Hispanic ethnicity. The propensity score model is shown in Supplemental Table S2. There were similar rates of cross-over between intervention and control groups: nearly 60% of the patients from both arms never changed units and the distribution of unit changes were similar between groups (Table 2).

**Table 2.** Univariate Study Outcomes.

|  | **Intervention**<br>**(Received alert)**<br>**(N=1506)** | **Control**<br>**(No alert)**<br>**(N=1274)** | **Difference [95%CI]** |
| --- | --- | --- | --- |
| **Type of Alerts during Admission** |  |  |  |
| Primary only | 930 (61.8%) | 746 (58.6%) | 3.2% [-0.5%, 6.9%] |
| RRT only | 250 (16.6%) | 243 (19.1%) | -2.5% [-5.4%, 0.5%] |
| Both Primary and RRT | 218 (14.5%) | 179 (14.1%) | 0.4% [-2.3%, 3.1%] |
| No alert | 108 (7.2%) | 106 (8.3%) | -1.1% [-3.2%, 0.9%] |
| **Number of RRT Alerts during Admission** |  |  |  |
| 0 | 1038 (68.9%) | 852 (66.9%) | 2.0% [-1.5%, 5.6%] |
| 1 | 376 (25.0%) | 350 (27.5%) | -2.5% [-5.9%, 0.9%] |
| 2 | 85 (5.6%) | 68 (5.3%) | 0.3% [-1.5%, 2.1%] |
| 3 | 7 (0.5%) | 4 (0.3%) | 0.2% [-0.4%, 0.7%] |
| **Number of Primary Alerts during Admission** |  |  |  |
| 0 | 358 (23.8%) | 349 (27.4%) | -3.6% [-7.0%, -0.3%] |
| 1 | 1148 (76.2%) | 925 (72.6%) |  |
| **Number of unit transfers during admission** |  |  |  |
| 0 | 868 (57.6%) | 762 (59.8%) | -2.2% [-5.9%, 1.6%] |
| 1 | 403 (26.8%) | 330 (25.9%) | 0.9% [-2.5%, 4.2%] |
| 2 | 145 (9.6%) | 107 (8.4%) | 1.2% [-1.0%, 3.4%] |
| 3+ | 90 (6.0%) | 75 (5.9%) | 0.1% [-1.7%, 1.9%] |
| **Escalation to stepdown or ICU** |  |  |  |
| Yes | 168 (11.2%) | 124 (9.7%) | 1.4% [-0.9%, 3.8%] |
| No | 1338 (88.8%) | 1150 (90.3%) |  |
| **Escalation to ICU only** |  |  |  |
| Yes | 115 (7.6%) | 100 (7.8%) | -0.2% [-2.3%, 1.9%] |
| No | 1391 (92.4%) | 1174 (92.2%) |  |
| **Escalation to ICU within 12 Hours** |  |  |  |
| Yes | 24 (1.6%) | 7 (0.5%) | 1.0% [0.2%, 1.9%] |
| No | 1482 (98.4%) | 1267 (99.5%) |  |
| **Escalation to ICU within 24 Hours** |  |  |  |
| Yes | 30 (2.0%) | 11 (0.9%) | 1.1% [0.2%, 2.1%] |
| No | 1476 (98.0%) | 1263 (99.1%) |  |
| **Time between Alert and escalation, hours** |  |  |  |
| Median [Q1, Q3] | 50.6 [21.6, 103] | 58.6 [25.4, 115] | -5.70 [-10.00, -2.00] |
| **Any order within six hours of an alert\*** |  |  |  |
| Yes | 907 (60.2%) | 747 (58.6%) | 1.6% (-2.1%, 5.3%) |
| No |  |  |  |

|  | Intervention<br>(Received alert)<br>(N=1506) | Control<br>(No alert)<br>(N=1274) | Difference [95%CI] |
| --- | --- | --- | --- |
| **Medication order within six hours of an alert** |  |  |  |
| Yes | 254 (16.9%) | 170 (13.3%) | 3.5% (0.8%, 6.3%) |
| No | 1252 (83.1%) | 1104 (86.7%) |  |
| **Cardiovascular support ordered within six hours of an alert\*\*** |  |  |  |
| Yes | 240 (15.9%) | 148 (11.6%) | 4.3% (1.7%, 6.9%) |
| No | 1266 (84.1%) | 1126 (88.4%) |  |
| **Last deterioration score before discharge** |  |  |  |
| Mean (SD) | 0.57 (0.13) | 0.58 (0.12) | 0.00 [0.00, 0.00] |
| **Hospital length of stay, days** |  |  |  |
| Median [Q1, Q3] | 5.77 [3.08, 10.7] | 6.02 [3.40, 12.0] | -0.31 [-1.00, 0.00] |
| **In-hospital mortality** |  |  |  |
| Yes | 81 (5.4%) | 84 (6.6%) | -1.2% [-3.1%, 0.6%] |
| No | 1425 (94.6%) | 1190 (93.4%) |  |
| **30-day mortality** |  |  |  |
| Yes | 87 (5.8%) | 100 (7.8%) | -2.1% [-4.0%, -0.1%] |
| No | 1419 (94.2%) | 1174 (92.2%) |  |
| **Combined in hospital and 30-day mortality** |  |  |  |
| Yes | 104 (6.9%) | 120 (9.4%) | -2.5% (-4.6%, -0.4%) |
| No | 1402 (93.1%) | 1154 (90.6%) |  |

\*Includes medications, fluids, ECG, laboratory, imaging and procedure orders
\*\*Includes Autonomic, Cardiovascular, Cardiac, and Diuretic medications, plus fluid bolus

### Primary Outcome

#### Escalations in Care

The incidence of escalation to a higher level of care in the intervention arm was 168/1,506 (11.2%) versus 124/1,274 (9.7%) in the control arm (absolute difference 1.4%; 95% CI -0.9% to 3.8%, p=0.25). The incidence of ICU only escalation in the intervention arm was 115/1,506 (7.6%) versus 100/1,274 (7.8%) in the control arm (absolute difference -0.2%; 95% CI -2.3% to 1.9%, p=0.89). In secondary analyses (Supplemental Tables S3-S6), we found no evidence that patient presence on individual units, the occurrence of any alert or the alert level (primary team versus RRT) influenced these results. IPTW log binomial regression modeling showed a relative risk of 1.22 (95% CI 0.97 to 1.54, p = 0.10) for escalation in the intervention group compared to controls. (Figure 2a)

**Figure 2:**
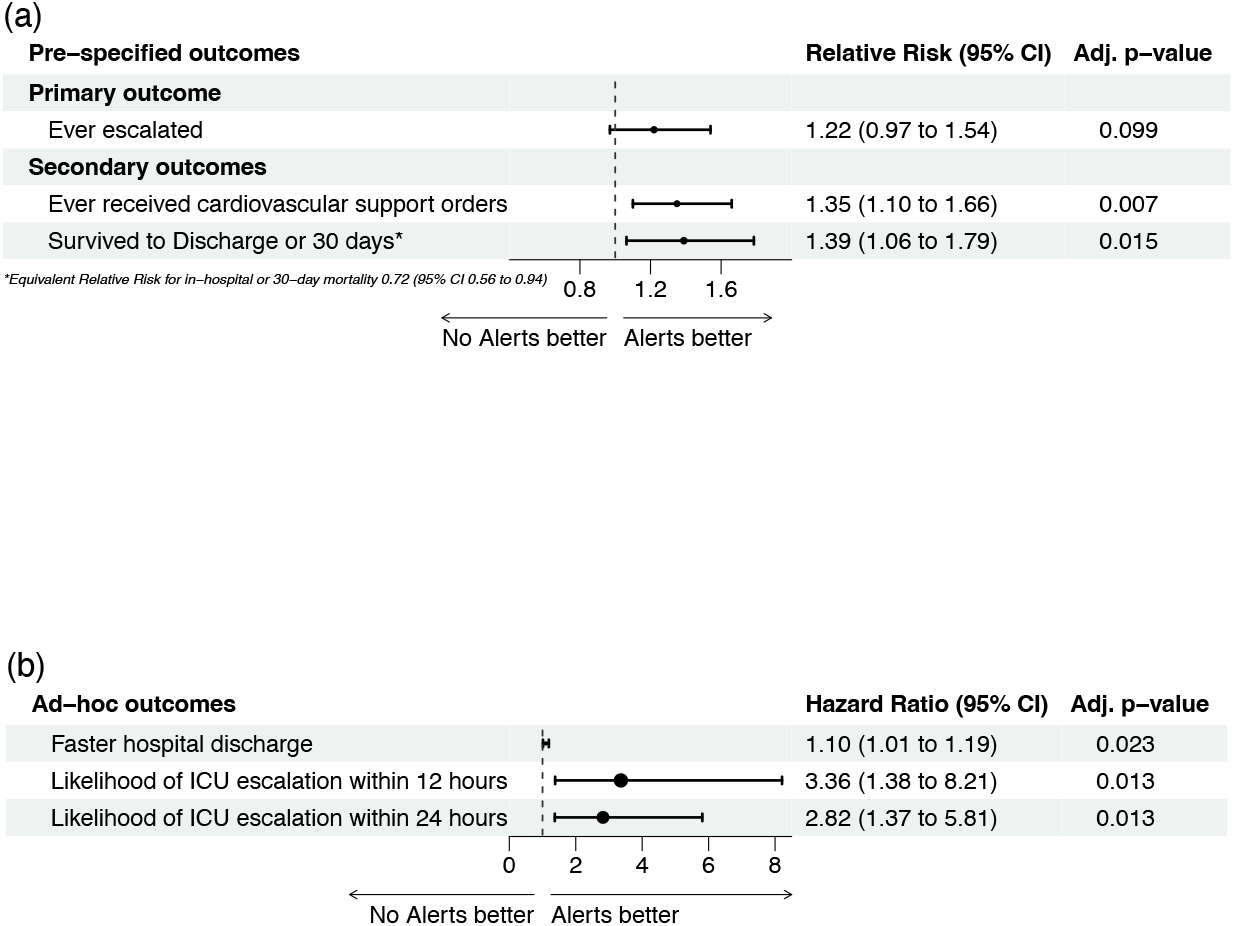
Forest Plots of Relative Risk and Hazard Ratio for pre-specified and ad-hoc outcomes. Forest plots showing the results of the IPTW adjusted analysis for the primary, secondary, and ad-hoc outcomes. The dot is the point estimate, and the bars shows the 95% confidence interval. Panel (a) shows the Relative Risk (RR) for the pre-specified primary and secondary outcomes. A RR > 1 indicates that alerts were associated with an improved outcome. “Survived to discharge or 30 days” is the inverse of the RR for combined in-hospital and 30-day mortality. Panel (b) shows the Hazard Ratio (HR) for the ad-hoc outcomes. A HR > 1 indicates that alerts were associated with an improved outcome. The p-values for the secondary outcomes and ad-hoc outcomes were adjusted using the false discovery rate method.

### Secondary Outcomes

#### Provider Orders

While the overall rate of orders placed within six hours of an alert were similar between groups (Table 2), more patients in the intervention arm received medication orders 254/1,506(16.9%) than in the control arm 170/1,274(13.3%) (absolute difference 3.5%;95% CI 0.8-6.3%, p=0.012). The difference in medication orders was primarily driven by orders for CV support, 240/1506 (15.9%) versus 148/1274 (11.6%) patients (absolute difference 4.3%;95% CI 1.7%-6.9%, p=0.001 (Table 2). An ECG was ordered for 128/1,506(8.5%) patients in the intervention arm versus 66/1,274(5.2%) patients in control arm (absolute difference 3.3%;95% CI 1.4-5.3%, p<0.001) (Supplemental Table S7). The IPTW negative binomial regression models confirmed that patients in the intervention group were more likely to have CV support orders placed (relative risk 1.35;95% CI 1.1-1.66, p=0.007). (Figure 2a)

#### In-hospital and 30-day Mortality

The combined in-hospital and 30-day mortality was significantly lower among patients who received an alert, 104/1,506 (6.9%) versus 120/1,274 (9.4%) (absolute difference -2.5%; 95% CI -4.6% to -0.4%, p=0.04). This result was confirmed by the IPTW log-binomial regression model (relative risk 0.72; 95% CI 0.56-0.94, p=0.015) (Figure 2a).

#### Alert Performance

The alerts performed at the patient level as expected in predicting escalation to ICU or stepdown. Sensitivity at the selected thresholds of 0.59 for the primary alert and 0.65 for the secondary alert was high for both primary (0.93) and RRT (0.88) alerts.

Consistent with the alert notification protocol, specificity was lower for the primary alerts (0.11) compared to the RRT alerts (0.33). The PPV was 0.07 for primary team alerts and 0.15 for RRT alerts. If a patient got both alerts, the sensitivity improved to 0.39 and the PPV to 0.27. NPV was high for all alerts (0.93-0.95). Full alert performance is shown in Supplemental Table S8.

### Ad-hoc Outcomes

#### Time to Escalation

The time to escalation was significantly shorter among intervention patients than controls. Patients on the intervention units escalated after a median of 50.6 [21.6 - 103] hours versus 58.6 [25.4 - 115] hours for control patients (median difference 5.7 hours; 95% CI -10.0 to -2.0, p=0.002). The percentage of patients in the intervention arm who were escalated to the ICU within 12 hours after an RRT alert was 24/1,506 (1.6%) versus 7/1,274 (0.5%) in the control arm (absolute difference 1.0%;95% CI 0.2-1.9%, p=0.02). The difference in escalation to the ICU within 24 hours of an alert was similar (Table 2). The IPTW Cox regression confirmed that intervention patients were more likely to go to the ICU within 12 hours (HR 1.38;95% CI 1.38-8.21, p=0.1) or 24 hours (HR 2.82;95% CI 1.37-5.81, p=0.01). (Figure 2b)

#### Length of Stay and Likelihood of Earlier Discharge

Patients assigned to the intervention units had both a shorter length of hospital stay, 5.8 [3.1-10.7] versus 6.0 [3.4-12] days (absolute difference -0.31 days; 95% CI 1.00-0.00, p=0.04) (Table 2) and a shorter unit stay (Supplemental Tables S3 and S4). The hazard ratio for faster hospital discharge in the IPTW Cox model was 1.10 (95% CI 1.01-1.19, p=0.023). (Figure 2b)

## Discussion

In this single-center pragmatic trial, the primary hypothesis that real-time alerts would prevent escalation of care was not confirmed. There was a non-significant trend towards increased escalation to stepdown or ICU in the intervention arm, and no significant difference in the rate of escalation to ICU only. Patients who received alerts, however, were significantly more likely to be transferred to an ICU within 12-24 hours of the alert. Alerts were also associated with more medication orders overall, and specifically more orders for cardiovascular support medications. Finally, alerts had statistically significant associations with shorter length of hospital stay and reduced mortality.

This investigation provides preliminary confirmation of the association of automated alerts with decreased length of stay and 30-day mortality described by several recent pre-post intervention studies. Escobar et al., studied the staggered deployment of automated alerts across a large 21 hospital health system and found that the relative risk for 30-day mortality among patients getting an alert was 0.84 (95% CI 0.78-0.9) compared to patients not receiving the alerts.^20^ A similar study by Winslow et al., conducted across a four-hospital mixed community/academic health system, found a significant decrease in all cause in-hospital mortality during the study period, from 13.9% to 8.8% among intermediate and high risk patients.^22^

In contrast, several older randomized controlled trials of automated alerts reported by Bailey et al. found no difference in either the proportion of patients transferred to the ICU or in-hospital mortality between groups, although hospital length of stay was statistically shorter in the intervention group.^8,11^ Possible methodological differences that explain the different results include lack of direct RRT team activation and limited involvement of the primary nursing and RRT teams in study design and implementation.

All these prior studies used logistic regression models. In contrast, we utilized a random forest model, which may perform better for high-dimensional data sets.^26,27^ Other investigators have likewise found random forest or gradient boosting machines to be the most accurate modeling techniques for predicting in-hospital mortality or ICU transfer after RRT evaluation.^13,28,29^ Our model also performed significantly better than a deep recurrent neural network trained on a similar set of features including hourly vitals.^30^ These findings suggest that random forest may be the “algorithm of choice” for real-time deterioration scores.

A key issue highlighted in all studies of automated clinical deterioration alerts is alarm fatigue for front-line staff that respond to the alerts.^31–33^ Bedoya et al reported that National Early Warning Score best practice alerts were “largely ignored by frontline nursing staff” in a retrospective cohort study that found no improvement in patient outcome metrics.^34^ We believe that the stratification of lower level alerts to primary nurses and higher level alerts to the RRT in the current investigation was effective in limiting alarm fatigue.

The positive predictive value of an RRT alert for escalation to intermediate or ICU level care was 15%, meaning that approximately one in six patients seen by RRT were escalated to a higher level of care after the alert. The word cloud shown in Figure 3 illustrates the most frequent replies of the RRT providers to the automated alert.

**Figure 3:**
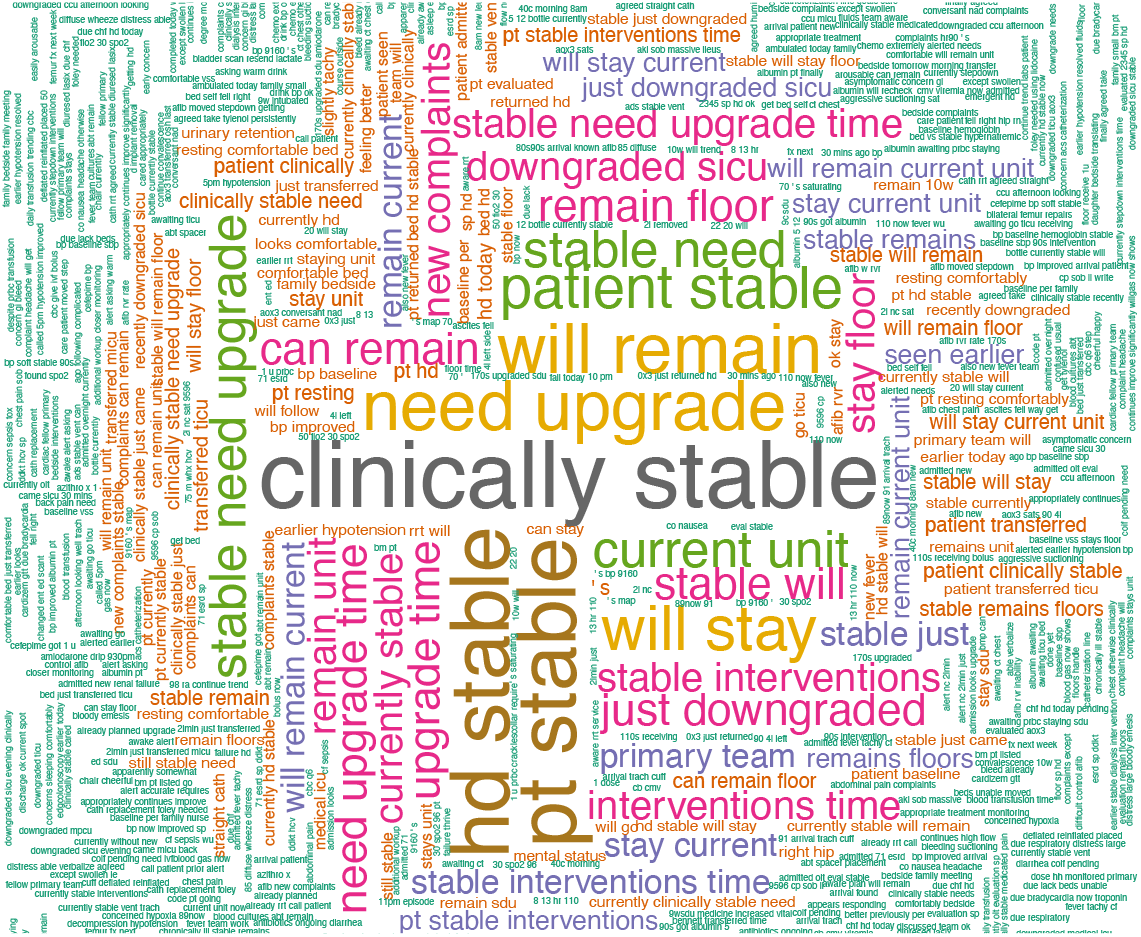
Provider Responses to Real-time Alerts. Word cloud showing the most frequently used phrases in the responses of the RRT provider to a real-time alert. The size and location of the words indicates the relative frequency of the phrase in the replies, with the most common phrases largest and closest to the center of the cloud. Providers most often rated patients as stable, despite a more rapid subsequent escalation to stepdown or ICU. The content of the smaller phrases around the periphery indicates that many of the patients seen by the RRT did have significant medical issues at the time of evaluation.

Although many of the RRT evaluations assessed patients as stable, the increased vigilance and extra steps taken (e.g., diagnostic or medication orders and more rapid escalations in care) were nonetheless associated with better patient outcomes. This suggests that the machine learning alerts were effective in selecting patients for additional evaluation and treatment.

A limitation of the current investigation is the curtailment of enrollment related to the onset of the COVID-19 pandemic. Approximately one-third of projected patients were enrolled, limiting statistical power. The primary hypothesis that alerts would decrease escalations in care is therefore not fully addressed by the current report. This may also have been due to the lack of an explicit escalation prevention intervention in the study protocol. The complex business process rules employed also differed from the simple 6-hour look-back rule employed during the model’s derivation and validation. The observed performance of the real time alerts therefore may not be directly comparable to the historical performance. The non-significant trend towards more escalations in care and the significantly shorter times to escalation nonetheless suggest that early involvement of the RRT could be an important factor in the observed improvements in length of stay and combined in-hospital and 30-day mortality.

A strength of the current investigation is the use of IPTW for estimating exposure effect standardized to a pseudo-population that removes confounding in observational studies.^24^ The IPTW relies on building a logistic regression model to estimate the probability of the intervention exposure for each individual, and on using the inverse of the predicted probability as a weight in subsequent analyses. We believe this was effective in controlling for the variability between intervention and control unit patient characteristics.

In conclusion, preliminary evidence suggests that real-time automated alerts of clinical deterioration, when stratified according to risk level, were effective in reducing time to escalations in care, hospital length of stay and mortality in inpatients admitted to medical-surgical units.

## Data Availability

Will individual participant data be available (including data dictionaries)?
Yes
What data in particular will be shared?
Individual participant data that underlie the results reported in this article (text, tables, figures, and appendices) will be available, after deidentification. A data dictionary will be provided if requested.
What other documents will be available?
Study protocol, statistical analysis plan, and analytic code.
When will data be available (start and end dates)?
Beginning 3 months and ending 5 years following article publication.
With whom?
Researchers who provide a methodologically sound proposal.
For what types of analyses?
To achieve the aims in the approved proposal.
By what mechanism will data be made available?
Proposals should be directed to. To gain access, data requestors will need to sign a data access agreement. Data will be provided in a format compatible with major statistical programs, directly to the requestor.

## Data Availability

The datasets generated and analyzed during the current study are available from the corresponding author on reasonable request.

## Code Availability

The R code used for data analysis is available from the corresponding author on reasonable request.

## References

1. Subbe, C. P., Kruger, M., Rutherford, P. & Gemmel, L. Validation of a modified Early Warning Score in medical admissions. QJM 94, 521–526 (2001).

2. McGinley, A. & Pearse, R. M. A national early warning score for acutely ill patients. BMJ 345, e5310 (2012).

3. National Early Warning Score (NEWS) 2. RCP London http://www.rcplondon.ac.uk/projects/outputs/national-early-warning-score-news-2 (2017).

4. Alam, N. et al. The impact of the use of the Early Warning Score (EWS) on patient outcomes: a systematic review. Resuscitation 85, 587–594 (2014).

5. McNeill, G. & Bryden, D. Do either early warning systems or emergency response teams improve hospital patient survival? A systematic review. Resuscitation 84, 1652–1667 (2013).

6. Ludikhuize, J., Smorenburg, S. M., de Rooij, S. E. & de Jonge, E. Identification of deteriorating patients on general wards; measurement of vital parameters and potential effectiveness of the Modified Early Warning Score. J. Crit. Care 27, 424.e7–13 (2012).

7. Ludikhuize, J. et al. Standardized measurement of the Modified Early Warning Score results in enhanced implementation of a Rapid Response System: A quasi-experimental study. Resuscitation 85, 676–682 (2014).

8. Bailey, T. C. et al. A trial of a real-time alert for clinical deterioration in patients hospitalized on general medical wards. J. Hosp. Med. 8, 236–242 (2013).

9. Rothman, M. J., Rothman, S. I. & Beals, J., 4th. Development and validation of a continuous measure of patient condition using the Electronic Medical Record. J. Biomed. Inform. 46, 837–848 (2013).

10. Churpek, M. M. et al. Multicenter development and validation of a risk stratification tool for ward patients. Am. J. Respir. Crit. Care Med. 190, 649–655 (2014).

11. Kollef, M. H. et al. A randomized trial of real-time automated clinical deterioration alerts sent to a rapid response team. J. Hosp. Med. 9, 424–429 (2014).

12. Finlay, G. D., Rothman, M. J. & Smith, R. A. Measuring the modified early warning score and the Rothman index: advantages of utilizing the electronic medical record in an early warning system. J. Hosp. Med. 9, 116–119 (2014).

13. Churpek, M. M. et al. Multicenter Comparison of Machine Learning Methods and Conventional Regression for Predicting Clinical Deterioration on the Wards. Crit. Care Med. 44, 368–374 (2016).

14. Hu, S. B., Wong, D. J. L., Correa, A., Li, N. & Deng, J. C. Prediction of Clinical Deterioration in Hospitalized Adult Patients with Hematologic Malignancies Using a Neural Network Model. PLoS One 11, e0161401 (2016).

15. Kipnis, P. et al. Development and validation of an electronic medical record-based alert score for detection of inpatient deterioration outside the ICU. J. Biomed. Inform. 64, 10–19 (2016).

16. Green, M. et al. Comparison of the Between the Flags calling criteria to the MEWS, NEWS and the electronic Cardiac Arrest Risk Triage (eCART) score for the identification of deteriorating ward patients. Resuscitation 123, 86–91 (2018).

17. Rubin, J. et al. An ensemble boosting model for predicting transfer to the pediatric intensive care unit. Int. J. Med. Inform. 112, 15–20 (2018).

18. Bartkowiak, B. et al. Validating the Electronic Cardiac Arrest Risk Triage (eCART) Score for Risk Stratification of Surgical Inpatients in the Postoperative Setting: Retrospective Cohort Study. Ann. Surg. 269, 1059–1063 (2019).

19. Churpek, M. M. et al. Validation of Early Warning Scores at Two Long-Term Acute Care Hospitals. Crit. Care Med. 47, e962–e965 (2019).

20. Escobar, G. J. et al. Automated Identification of Adults at Risk for In-Hospital Clinical Deterioration. N. Engl. J. Med. 383, 1951–1960 (2020).

21. Romero-Brufau, S. et al. Using machine learning to improve the accuracy of patient deterioration predictions: Mayo Clinic Early Warning Score (MC-EWS). J. Am. Med. Inform. Assoc. 28, 1207–1215 (2021).

22. Winslow, C. J. et al. The Impact of a Machine Learning Early Warning Score on Hospital Mortality: A Multicenter Clinical Intervention Trial. Crit. Care Med. (2022) doi:10.1097/CCM.0000000000005492.

23. Kia, A. et al. MEWS++: Enhancing the Prediction of Clinical Deterioration in Admitted Patients through a Machine Learning Model. J. Clin. Med. Res. 9, (2020).

24. Mansournia, M. A. & Altman, D. G. Inverse probability weighting. BMJ 352, i189 (2016).

25. Benjamini, Y. & Hochberg, Y. Controlling the false discovery rate: A practical and powerful approach to multiple testing. J. R. Stat. Soc. 57, 289–300 (1995).

26. Couronné, R., Probst, P. & Boulesteix, A.-L. Random forest versus logistic regression: a large-scale benchmark experiment. BMC Bioinformatics 19, 270 (2018).

27. Touw, W. G. et al. Data mining in the Life Sciences with Random Forest: a walk in the park or lost in the jungle? Brief. Bioinform. 14, 315–326 (2013).

28. Shappell, C., Snyder, A., Edelson, D. P., Churpek, M. M. & American Heart Association’s Get With The Guidelines-Resuscitation Investigators. Predictors of In-Hospital Mortality After Rapid Response Team Calls in a 274 Hospital Nationwide Sample. Crit. Care Med. 46, 1041–1048 (2018).

29. Akel, M. A., Carey, K. A., Winslow, C. J., Churpek, M. M. & Edelson, D. P. Less is more: Detecting clinical deterioration in the hospital with machine learning using only age, heart rate, and respiratory rate. Resuscitation 168, 6–10 (2021).

30. Shah, P. K. et al. A Simulated Prospective Evaluation of a Deep Learning Model for Real-Time Prediction of Clinical Deterioration Among Ward Patients. Crit. Care Med. 49, 1312–1321 (2021).

31. Fleischman, W., Ciliberto, B., Rozanski, N., Parwani, V. & Bernstein, S. L. Emergency department monitor alarms rarely change clinical management: An observational study. Am. J. Emerg. Med. 38, 1072–1076 (2020).

32. Cvach, M. Monitor alarm fatigue: an integrative review. Biomed. Instrum. Technol. 46, 268–277 (2012).

33. Yiu, C. J., Khan, S. U., Subbe, C. P., Tofeec, K. & Madge, R. A. Into the night: factors affecting response to abnormal Early Warning Scores out-of-hours and implications for service improvement. Acute Med. 13, 56–60 (2014).

34. Bedoya, A. D. et al. Minimal Impact of Implemented Early Warning Score and Best Practice Alert for Patient Deterioration. Crit. Care Med. 1 (2018).

