## Supplementary Material for "Real-time Machine Learning Alerts to Prevent Escalation of Care: A Pragmatic Clinical Trial"

### Figure S1 – Process flow chart

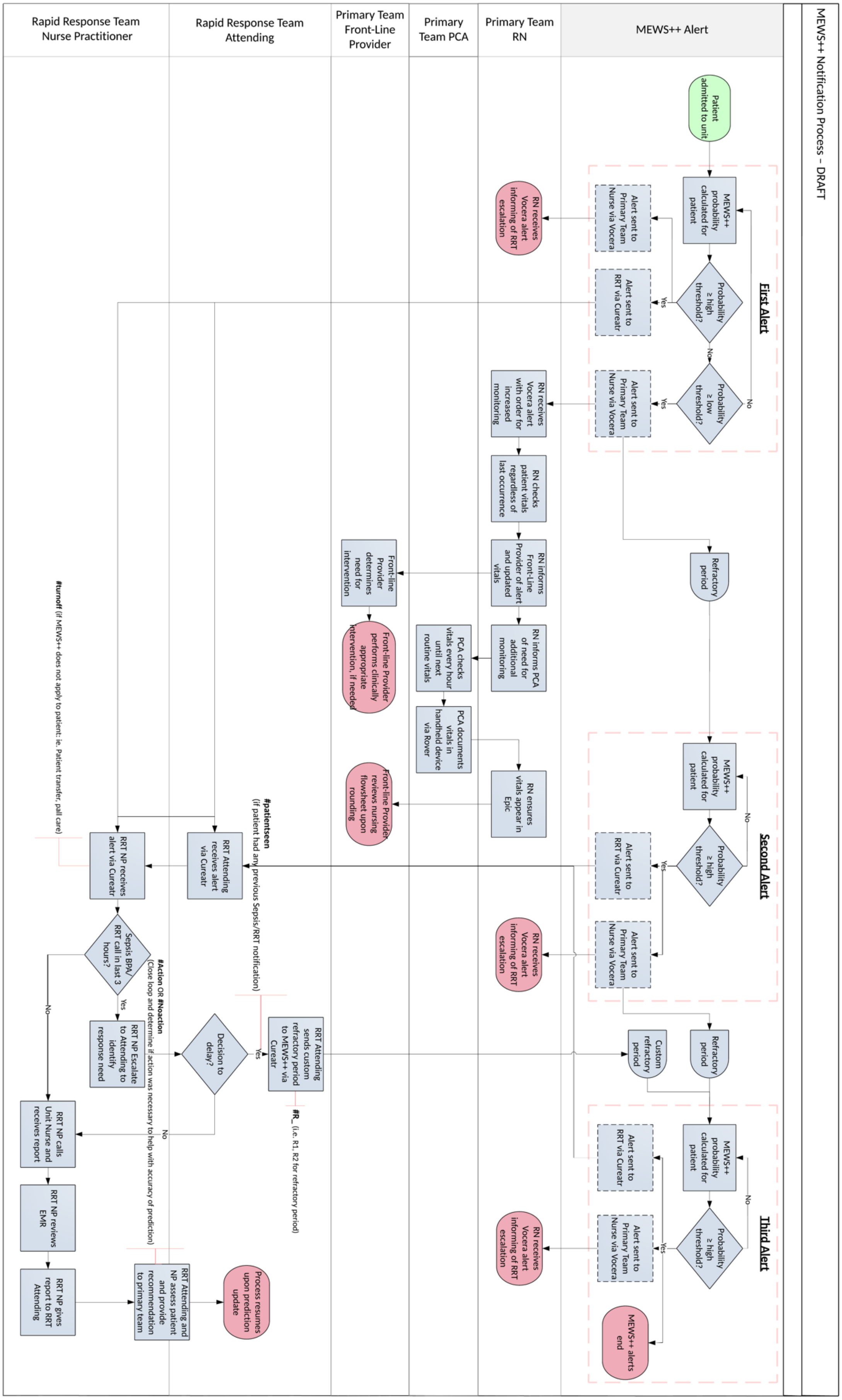

#### Table S1 - Group Comparison for Patients Missing Comorbidity Data

| | Total (N=2780) | Comorbidities Missing (N=190) | No Missing (N=2590) | Diff (95%CI) |
|---|---|---|---|---|
| **Age** | | | | |
| Mean (SD) | 66.3 (17.4) | 58.8 (17.7) | 66.9 (17.3) | -8.08 (-10.70, -5.47) |
| **Gender** | | | | |
| Female | 1446 (52.0%) | 99 (52.1%) | 1347 (52.0%) | 0.1% (-7.4%, 7.6%) |
| Male | 1334 (48.0%) | 91 (47.9%) | 1243 (48.0%) | |
| **Race** | | | | |
| White | 900 (34.2%) | 11 (27.5%) | 889 (34.3%) | -6.8% (-22.1%, 8.4%) |
| Black | 622 (23.7%) | 11 (27.5%) | 611 (23.6%) | 3.9% (-11.3%, 19.1%) |
| Asian | 172 (6.5%) | 5 (12.5%) | 167 (6.4%) | 6.1% (-5.5%, 17.6%) |
| Other | 936 (35.6%) | 13 (32.5%) | 923 (35.6%) | -3.1% (-19.0%, 12.8%) |
| **Ethnicity** | | | | |
| Not Hispanic or Latino | 1747 (66.4%) | 27 (67.5%) | 1720 (66.4%) | 1.1% (-14.6%, 16.8%) |
| Hispanic or Latino | 883 (33.6%) | 13 (32.5%) | 870 (33.6%) | |
| **BMI ($kg \cdot m^{-2}$)** | | | | |
| Mean (SD) | 26.9 (8.34) | 28.3 (5.67) | 26.9 (8.38) | 1.39 (-0.45, 3.23) |
| **BMI Category** | | | | |
| Normal | 961 (37.1%) | 6 (15.0%) | 955 (37.5%) | -3.6% (-11.7%, 4.5%) |
| Underweight | 222 (8.6%) | 2 (5.0%) | 220 (8.6%) | -22.5% (-35.0%, -10.0%) |
| Overweight | 717 (27.7%) | 15 (37.5%) | 702 (27.5%) | 10.0% (-6.4%, 26.3%) |
| Obese | 689 (26.6%) | 17 (42.5%) | 672 (26.4%) | 16.1% (-0.5%, 32.8%) |
| **In-hospital Mortality** | | | | |
| Yes | 165 (5.9%) | 1 (0.5%) | 164 (6.3%) | -5.8% (-7.5%, -4.1%) |
| No | 2615 (94.1%) | 189 (99.5%) | 2426 (93.7%) | |
| **30-day Mortality** | | | | |
| Yes | 187 (6.7%) | 2 (1.1%) | 185 (7.1%) | -6.1% (-8.1%, -4.1%) |
| No | 2593 (93.3%) | 188 (98.9%) | 2405 (92.9%) | |
| **Escalation to Stepdown or ICU** | | | | |
| Yes | 292 (10.5%) | 2 (1.1%) | 290 (11.2%) | -10.1% (-12.3%, -8.0%) |
| No | 2488 (89.5%) | 188 (98.9%) | 2300 (88.8%) | |
| **Escalation to ICU** | | | | |
| Yes | 215 (7.7%) | 0 (0%) | 215 (8.3%) | -8.3% (-9.6%, -7.0%) |
| No | 2565 (92.3%) | 190 (100%) | 2375 (91.7%) | |
| **Hospital Length of Stay** | | | | |
| Median [Q1, Q3] | 5.83 [3.22, 11.2] | 2.86 [1.63, 5.75] | 6.00 [3.60, 11.7] | -2.83 (-3.00, -2.00) |
| **First Deterioration Score on Admission** | | | | |
| Mean (SD) | 0.57 (0.098) | 0.51 (0.064) | 0.57 (0.099) | -0.07 (-0.08, -0.06) |

**Table S2 - Propensity Score Matching Logistic Model**

| *Predictors* | *Odds Ratios* | *std. Error* | *CI* | *p* |
|---|---|---|---|---|
| **Intercept** | 2.024 | 0.616 | 1.117 – 3.681 | **0.020** |
| **Gender (Male)** | 1.103 | 0.093 | 0.935 – 1.302 | 0.246 |
| **Age** | 1.006 | 0.003 | 1.001 – 1.012 | **0.027** |
| **Race** | | | | |
| Black | 1.004 | 0.114 | 0.803 – 1.255 | 0.974 |
| Asian | 0.551 | 0.096 | 0.391 – 0.774 | **0.001** |
| Other | 0.799 | 0.103 | 0.620 – 1.027 | 0.080 |
| **Hispanic or Latino** | 0.986 | 0.119 | 0.779 – 1.249 | 0.905 |
| **BMI Category** | | | | |
| Underweight | 0.973 | 0.147 | 0.723 – 1.309 | 0.855 |
| Overweight | 1.161 | 0.118 | 0.952 – 1.417 | 0.141 |
| Obese | 1.368 | 0.146 | 1.109 – 1.688 | **0.003** |
| **Alcohol Use** | 0.627 | 0.107 | 0.447 – 0.874 | **0.006** |
| **HIV** | 1.040 | 0.196 | 0.720 – 1.507 | 0.835 |
| **Arrhythmia** | 0.851 | 0.164 | 0.583 – 1.241 | 0.400 |
| **Asthma/COPD** | 1.018 | 0.102 | 0.836 – 1.241 | 0.856 |
| **Dementia** | 0.887 | 0.139 | 0.653 – 1.207 | 0.445 |
| **Depression** | 0.963 | 0.122 | 0.752 – 1.234 | 0.765 |
| **Diabetes** | 0.895 | 0.082 | 0.748 – 1.072 | 0.230 |
| **Drug Abuse** | 1.014 | 0.201 | 0.687 – 1.498 | 0.946 |
| **Hypertension** | 1.067 | 0.104 | 0.881 – 1.293 | 0.506 |
| **Thyroid Disease** | 0.886 | 0.103 | 0.705 – 1.112 | 0.295 |
| **Prior Malignancy** | 1.362 | 0.131 | 1.129 – 1.645 | **0.001** |
| **Peripheral Vascular Disease** | 1.266 | 0.186 | 0.950 – 1.693 | 0.109 |
| **First Deterioration Score on Admission** | 0.837 | 0.035 | 0.770 – 0.908 | **<0.001** |
| **Missing Comorbidity Data** | 0.463 | 0.159 | 0.231 – 0.899 | **0.025** |

#### Table S3 - Unit Level Analysis – All unit admissions

| | Total (N=4625) | Intervention (Received alert) (N=2652) | Control (No alert) (N=1973) | Diff (95%CI) |
|---|---|---|---|---|
| **Type of Alerts in the Unit** | | | | |
| Primary only | 1773 (38.3%) | 1007 (38.0%) | 766 (38.8%) | -0.9% (-3.7%, 2.0%) |
| RRT only | 625 (13.5%) | 335 (12.6%) | 290 (14.7%) | -2.1% (-4.1%, 0.0%) |
| Both Primary and RRT | 300 (6.5%) | 158 (6.0%) | 142 (7.2%) | -1.2% (-2.7%, 0.3%) |
| No alert | 1927 (41.7%) | 1152 (43.4%) | 775 (39.3%) | 4.2% (1.3%, 7.1%) |
| **Number of RRT Alerts in the Unit** | | | | |
| 0 | 3700 (80.0%) | 2159 (81.4%) | 1541 (78.1%) | 3.3% (0.9%, 5.7%) |
| 1 | 790 (17.1%) | 418 (15.8%) | 372 (18.9%) | -3.1% (-5.4%, -0.8%) |
| 2 | 130 (2.8%) | 73 (2.8%) | 57 (2.9%) | -0.1% (-1.1%, 0.9%) |
| 3 | 5 (0.1%) | 2 (0.1%) | 3 (0.2%) | -0.1% (-0.3%, 0.2%) |
| **Number of Primary Alerts in the Unit** | | | | |
| 0 | 2552 (55.2%) | 1487 (56.1%) | 1065 (54.0%) | 2.1% (-0.9%, 5.0%) |
| 1 | 2073 (44.8%) | 1165 (43.9%) | 908 (46.0%) | |
| **Escalation to Stepdown or ICU** | | | | |
| Yes | 325 (7.0%) | 191 (7.2%) | 134 (6.8%) | 0.4% (-1.1%, 1.9%) |
| No | 4300 (93.0%) | 2461 (92.8%) | 1839 (93.2%) | |
| **Escalation to ICU only** | | | | |
| Yes | 234 (5.1%) | 129 (4.9%) | 105 (5.3%) | -0.5% (-1.8%, 0.9%) |
| No | 4391 (94.9%) | 2523 (95.1%) | 1868 (94.7%) | |
| **Escalation to ICU within 12 Hours** | | | | |
| Yes | 31 (0.7%) | 24 (0.9%) | 7 (0.4%) | 0.6% (0.1%, 1.0%) |
| No | 4594 (99.3%) | 2628 (99.1%) | 1966 (99.6%) | |
| **Escalation to ICU within 24 Hours** | | | | |
| Yes | 42 (0.9%) | 30 (1.1%) | 12 (0.6%) | 0.5% (-0.1%, 1.1%) |
| No | 4583 (99.1%) | 2622 (98.9%) | 1961 (99.4%) | |
| **Time between Alert and ICU admission, hours** | | | | |
| Median [Q1, Q3] | 52.9 [21.9, 109] | 48.6 [19.8, 103] | 58.3 [24.4, 116] | -6.70 (-11.00, -3.00) |
| **Any order within six hours of an alert** | | | | |
| Yes | 1716 (37.1%) | 963 (36.3%) | 753 (38.2%) | -1.9% (-4.7%, 1.0%) |
| No | 2909 (62.9%) | 1689 (63.7%) | 1220 (61.8%) | |
| **Number of orders within six hours** | | | | |
| Median [Q1, Q3] | 5.00 [2.00, 8.00] | 5.00 [2.00, 8.00] | 5.00 [2.00, 9.00] | 0.00 (-1.00, 0.00) |
| **Medication order within six hours of an alert** | | | | |
| Yes | 435 (9.4%) | 264 (10.0%) | 171 (8.7%) | 1.3% (-0.4%, 3.0%) |
| No | 4190 (90.6%) | 2388 (90.0%) | 1802 (91.3%) | |
| **Number of medication orders** | | | | |
| Median [Q1, Q3] | 1.00 [1.00, 2.00] | 1.00 [1.00, 2.00] | 1.00 [1.00, 2.00] | 0.00 (0.00, 0.00) |
| **Unit Length of Stay, days** | | | | |
| Median [Q1, Q3] | 2.88 [1.43, 5.53] | 2.79 [1.28, 5.09] | 3.04 [1.58, 5.76] | -0.24 (0.00, 0.00) |

#### Table S4 - Unit Level Analysis – First unit admission

Unit Level - First Unit Admission

| | Total (N=2780) | Intervention (Received alert) (N=1506) | Control (No alert) (N=1274) | Diff (95%CI) |
|---|---|---|---|---|
| **Type of Alerts in the Unit** | | | | |
| Primary only | 1561 (56.2%) | 862 (57.2%) | 699 (54.9%) | 2.4% (-1.4%, 6.1%) |
| RRT only | 467 (16.8%) | 237 (15.7%) | 230 (18.1%) | -2.3% (-5.2%, 0.6%) |
| Both Primary and RRT | 288 (10.4%) | 151 (10.0%) | 137 (10.8%) | -0.7% (-3.1%, 1.6%) |
| No alert | 464 (16.7%) | 256 (17.0%) | 208 (16.3%) | 0.7% (-2.2%, 3.5%) |
| **Number of RRT Alerts in the Unit** | | | | |
| 0 | 2025 (72.8%) | 1118 (74.2%) | 907 (71.2%) | 3.0% (-0.4%, 6.4%) |
| 1 | 641 (23.1%) | 325 (21.6%) | 316 (24.8%) | -3.2% (-6.4%, 0.0%) |
| 2 | 109 (3.9%) | 61 (4.1%) | 48 (3.8%) | 0.3% (-1.2%, 1.8%) |
| 3 | 5 (0.2%) | 2 (0.1%) | 3 (0.2%) | -0.1% (-0.5%, 0.3%) |
| **Number of Primary Alerts in the Unit** | | | | |
| 0 | 931 (33.5%) | 493 (32.7%) | 438 (34.4%) | -1.6% (-5.2%, 2.0%) |
| 1 | 1849 (66.5%) | 1013 (67.3%) | 836 (65.6%) | |
| **Escalation to Stepdown or ICU** | | | | |
| Yes | 200 (7.2%) | 113 (7.5%) | 87 (6.8%) | 0.7% (-1.3%, 2.7%) |
| No | 2580 (92.8%) | 1393 (92.5%) | 1187 (93.2%) | |
| **Escalation to ICU only** | | | | |
| Yes | 142 (5.1%) | 76 (5.0%) | 66 (5.2%) | -0.1% (-1.9%, 1.6%) |
| No | 2638 (94.9%) | 1430 (95.0%) | 1208 (94.8%) | |
| **Escalation to ICU within 12 Hours** | | | | |
| No | 2752 (99.0%) | 1484 (98.5%) | 1268 (99.5%) | -1.0% (-1.8%, -0.2%) |
| Yes | 28 (1.0%) | 22 (1.5%) | 6 (0.5%) | |
| **Escalation to ICU within 24 Hours** | | | | |
| No | 2743 (98.7%) | 1479 (98.2%) | 1264 (99.2%) | -1.0% (-1.9%, -0.1%) |
| Yes | 37 (1.3%) | 27 (1.8%) | 10 (0.8%) | |
| **Time between Alert and ICU admission, hours** | | | | |
| Median [Q1, Q3] | 51.1 [21.7, 108] | 46.8 [19.4, 100] | 57.2 [24.1, 115] | -7.15 (-11.00, -3.00) |
| **Any order within six hours of an alert** | | | | |
| No | 1306 (47.0%) | 702 (46.6%) | 604 (47.4%) | -0.8% (-4.6%, 3.0%) |
| Yes | 1474 (53.0%) | 804 (53.4%) | 670 (52.6%) | |
| **Number of orders within six hours** | | | | |
| Median [Q1, Q3] | 1.00 [0, 5.00] | 1.00 [0, 5.00] | 1.00 [0, 6.00] | 0.00 (0.00, 0.00) |
| **Medication order within six hours of an alert** | | | | |
| No | 2396 (86.2%) | 1274 (84.6%) | 1122 (88.1%) | -3.5% (-6.1%, -0.9%) |
| Yes | 384 (13.8%) | 232 (15.4%) | 152 (11.9%) | |
| **Number of medication orders** | | | | |
| Median [Q1, Q3] | 0 [0, 0] | 0 [0, 0] | 0 [0, 0] | 0.00 (0.00, 0.00) |
| **Unit Length of Stay, days** | | | | |
| Median [Q1, Q3] | 2.77 [1.33, 5.01] | 2.66 [1.16, 4.80] | 2.90 [1.56, 5.32] | -0.27 (0.00, 0.00) |

#### Table S5 - Alert Level Analysis – All Alerts

| | Total (N=3138) | Intervention (Received alert) (N=1735) | Control (No alert) (N=1403) | Diff (95%CI) |
|---|---|---|---|---|
| **Escalation to ICU after alert** | | | | |
| Yes | 203 (6.5%) | 124 (7.1%) | 79 (5.6%) | 1.5% (-0.3%, 3.3%) |
| No | 2935 (93.5%) | 1611 (92.9%) | 1324 (94.4%) | |
| **Escalation to ICU within 12 Hours** | | | | |
| Yes | 60 (1.9%) | 43 (2.5%) | 17 (1.2%) | 1.3% (0.3%, 2.3%) |
| No | 3078 (98.1%) | 1692 (97.5%) | 1386 (98.8%) | |
| **Escalation to ICU within 24 Hours** | | | | |
| Yes | 92 (2.9%) | 63 (3.6%) | 29 (2.1%) | 1.6% (0.3%, 2.8%) |
| No | 3046 (97.1%) | 1672 (96.4%) | 1374 (97.9%) | |
| **Time between alert and ICU admission, hours** | | | | |
| Median [Q1, Q3] | 55.7 [22.3, 116] | 52.1 [20.0, 111] | 63.1 [25.5, 122] | -7.40 (-11.00, -4.00) |
| **Any order within six hours of an alert** | | | | |
| Yes | 1922 (61.2%) | 1073 (61.8%) | 849 (60.5%) | 1.3% (-2.2%, 4.8%) |
| No | 1216 (38.8%) | 662 (38.2%) | 554 (39.5%) | |
| **Number of orders within six hours** | | | | |
| Median [Q1, Q3] | 4 [2, 7] | 4 [2, 7] | 5 [2, 8] | 0.00 (-1.00, 0.00) |
| **Medication orders within six hours** | | | | |
| Yes | 457 (14.6%) | 280 (16.1%) | 177 (12.6%) | 3.5% (1.0%, 6.0%) |
| No | 2681 (85.4%) | 1455 (83.9%) | 1226 (87.4%) | |
| **Cardiovascular medication orders within six hours** | | | | |
| Yes | 102 (3.3%) | 68 (3.9%) | 34 (2.4%) | 1.5% (0.2%, 2.8%) |
| No | 3036 (96.7%) | 1667 (96.1%) | 1369 (97.6%) | |
| **Imaging, Fluid, Laboratory, ECG, or Respiratory Therapy Orders** | | | | |
| Yes | 1802 (57.4%) | 999 (57.6%) | 803 (57.2%) | 0.3% (-3.2%, 3.9%) |
| No | 1336 (42.6%) | 736 (42.4%) | 600 (42.8%) | |

#### Table S6 - Alert Level Analysis, RRT Alerts only

| | Total (N=1065) | Intervention (Received alert) (N=570) | Control (No alert) (N=495) | Diff (95%CI) |
|---|---|---|---|---|
| **Escalation to ICU after alert** | | | | |
| Yes | 116 (10.9%) | 68 (11.9%) | 48 (9.7%) | 2.2% (-1.7%, 6.1%) |
| No | 949 (89.1%) | 502 (88.1%) | 447 (90.3%) | |
| **Escalation to ICU within 12 Hours** | | | | |
| Yes | 31 (2.9%) | 24 (4.2%) | 7 (1.4%) | 2.8% (0.7%, 4.9%) |
| No | 1034 (97.1%) | 546 (95.8%) | 488 (98.6%) | |
| **Escalation to ICU within 24 Hours** | | | | |
| Yes | 45 (4.2%) | 32 (5.6%) | 13 (2.6%) | 3.0% (0.4%, 5.5%) |
| No | 1020 (95.8%) | 538 (94.4%) | 482 (97.4%) | |
| **Time between alert and ICU admission, hours** | | | | |
| Median [Q1, Q3] | 68.8 [27.4, 139] | 63.1 [24.5, 130] | 74.0 [31.4, 145] | -8.90 (-17.00, -2.00) |
| **Any order within six hours of an alert** | | | | |
| Yes | 688 (64.6%) | 378 (66.3%) | 310 (62.6%) | 3.7% (-2.3%, 9.6%) |
| No | 377 (35.4%) | 192 (33.7%) | 185 (37.4%) | |
| **Number of orders within six hours** | | | | |
| Median [Q1, Q3] | 5 [2, 7] | 5 [2, 8] | 5 [2, 7] | 0.00 (0.00, 0.00) |
| **Medication orders within six hours** | | | | |
| Yes | 169 (15.9%) | 101 (17.7%) | 68 (13.7%) | 4.0% (-0.6%, 8.5%) |
| No | 896 (84.1%) | 469 (82.3%) | 427 (86.3%) | |
| **Cardiovascular medication orders within six hours** | | | | |
| Yes | 47 (4.4%) | 28 (4.9%) | 19 (3.8%) | 1.1% (-1.6%, 3.7%) |
| No | 1018 (95.6%) | 542 (95.1%) | 476 (96.2%) | |
| **Imaging, Fluid, Laboratory,ECG, or Respiratory Therapy Orders** | | | | |
| Yes | 641 (60.2%) | 351 (61.6%) | 290 (58.6%) | 3.0% (-3.1%, 9.1%) |
| No | 424 (39.8%) | 219 (38.4%) | 205 (41.4%) | |

#### Table S7 – Patient orders placed within six hours of alert, by type

| | Total (N=2780) | Intervention (Received alert) (N=1506) | Control (No alert) (N=1274) | Diff (95%CI) |
|---|---|---|---|---|
| **Any medication ordered** | | | | |
| Yes | 424 (15.3%) | 254 (16.9%) | 170 (13.3%) | 3.5% (0.8%, 6.3%) |
| No | 2356 (84.7%) | 1252 (83.1%) | 1104 (86.7%) | |
| **Number of medication orders** | | | | |
| Median [Q1, Q3] | 1 [1, 2] | 1 [1, 2] | 1 [1, 2] | 0.00 (0.00, 0.00) |
| **Antihyperglycemic drugs ordered*** | | | | |
| Yes | 38 (1.4%) | 23 (1.5%) | 15 (1.2%) | 0.3% (-0.6%, 1.3%) |
| No | 2742 (98.6%) | 1483 (98.5%) | 1259 (98.8%) | |
| **Autonomic drugs ordered*** | | | | |
| Yes | 2 (0.1%) | 2 (0.1%) | 0 (0%) | 0.1% (-0.1%, 0.4%) |
| No | 2778 (99.9%) | 1504 (99.9%) | 1274 (100%) | |
| **Cardiovascular drugs ordered*** | | | | |
| Yes | 38 (1.4%) | 27 (1.8%) | 11 (0.9%) | 0.9% (0.0%, 1.8%) |
| No | 2742 (98.6%) | 1479 (98.2%) | 1263 (99.1%) | |
| **Cardiac drugs ordered*** | | | | |
| Yes | 9 (0.3%) | 7 (0.5%) | 2 (0.2%) | 0.3% (-0.2%, 0.8%) |
| No | 2771 (99.7%) | 1499 (99.5%) | 1272 (99.8%) | |
| **Diuretics ordered*** | | | | |
| Yes | 53 (1.9%) | 35 (2.3%) | 18 (1.4%) | 0.9% (-0.2%, 2.0%) |
| No | 2727 (98.1%) | 1471 (97.7%) | 1256 (98.6%) | |
| **Intravenous fluid/albumin ordered** | | | | |
| Yes | 349 (12.6%) | 202 (13.4%) | 147 (11.5%) | 1.9% (-0.7%, 4.4%) |
| No | 2431 (87.4%) | 1304 (86.6%) | 1127 (88.5%) | |
| **Any cardiovascular medication ordered**** | | | | |
| Yes | 388 (14.0%) | 240 (15.9%) | 148 (11.6%) | 4.3% (1.7%, 6.9%) |
| No | 2682 (96.5%) | 1438 (95.5%) | 1244 (97.6%) | |
| **Blood products ordered** | | | | |
| Yes | 286 (10.3%) | 151 (10.0%) | 135 (10.6%) | -0.6% (-2.9%, 1.8%) |
| No | 494 (89.7%) | 1355 (90.0%) | 1139 (89.4%) | |
| **ECG ordered** | | | | |
| Yes | 186 (6.7%) | 123 (8.2%) | 63 (4.9%) | 3.2% (1.3%, 5.1%) |

| | Total (N=2780) | Intervention (Received alert) (N=1506) | Control (No alert) (N=1274) | Diff (95%CI) |
|---|---|---|---|---|
| No | 2594 (93.3%) | 1383 (91.8%) | 1211 (95.1%) | |
| **Laboratory test ordered** | | | | |
| Yes | 1258 (45.3%) | 694 (46.1%) | 564 (44.3%) | 1.8% (-2.0%, 5.6%) |
| No | 1522 (54.7%) | 812 (53.9%) | 710 (55.7%) | |
| **Point of care testing ordered** | | | | |
| Yes | 377 (13.6%) | 217 (14.4%) | 160 (12.6%) | 1.9% (-0.8%, 4.5%) |
| No | 2403 (86.4%) | 1289 (85.6%) | 1114 (87.4%) | |
| **Imaging ordered** | | | | |
| Yes | 299 (10.8%) | 165 (11.0%) | 134 (10.5%) | 0.4% (-1.9%, 2.8%) |
| No | 2481 (89.2%) | 1341 (89.0%) | 1140 (89.5%) | |
| **Procedures ordered** | | | | |
| Yes | 21 (0.8%) | 14 (0.9%) | 7 (0.5%) | 0.4% (-0.3%, 1.1%) |
| No | 2759 (99.2%) | 1492 (99.1%) | 1267 (99.5%) | |
| **Respiratory Therapy ordered** | | | | |
| Yes | 20 (0.7%) | 15 (1.0%) | 5 (0.4%) | 0.6% (-0.1%, 1.3%) |
| No | 2760 (99.3%) | 1491 (99.0%) | 1269 (99.6%) | |
| **Any diagnostic test, therapy, or procedure ordered*** ** | | | | |
| Yes | 1440 (51.8%) | 794 (52.7%) | 646 (50.7%) | 2.0% (-1.8%, 5.8%) |
| No | 1340 (48.2%) | 712 (47.3%) | 628 (49.3%) | |

*Pharmaceutical class
**Includes Autonomic, Cardiac, Cardiovascular and Diuretic classes, plus Fluid orders
***Includes Imaging, Laboratory, ECG, and Respiratory Therapy Orders

**Table S8 - Alert Performance**

| | Accuracy | Sensitivity | Specificity | PPV* | NPV** |
|---|---|---|---|---|---|
| **Patient Level** | | | | | |
| Ever RRT (vs. never RRT) -Escalated | 0.71 [0.68 – 0.73] | 0.57 | 0.72 | 0.21 | 0.93 |
| Any alert (vs. No alert) - Escalated | 0.18 [0.16 – 0.2] | 0.97 | 0.08 | 0.12 | 0.95 |
| RRT only (vs. No Alert) - Escalated | 0.39 [ 0.34 – 0.45] | 0.88 | 0.33 | 0.15 | 0.95 |
| Primary only (vs. No Alert) - Escalated | 0.16 [ 0.14 – 0.19 | 0.93 | 0.11 | 0.07 | 0.95 |
| Both Alerts only (vs. No Alert) - Escalated | 0.49 [0.44 – 0.55] | 0.92 | 0.39 | 0.27 | 0.95 |
| Ever RRT (vs. never RRT) - ICU | 0.71 [0.69 – 0.73] | 0.65 | 0.72 | 0.16 | 0.96 |
| Any alert (vs. No alert) - ICU | 0.14 [0.13 – 0.16] | 0.97 | 0.08 | 0.08 | 0.97 |
| RRT only (vs. No Alert) - ICU | 0.39 [0.33 – 0.44] | 0.92 | 0.33 | 0.13 | 0.97 |
| Primary only (vs. No Alert) - ICU | 0.14 [0.12 – 0.16] | 0.92 | 0.11 | 0.04 | 0.97 |
| Both Alerts only (vs. No Alert) - ICU | 0.45 [0.39 – 0.5] | 0.93 | 0.37 | 0.19 | 0.97 |
| **Unit Level** | | | | | |
| Ever RRT (vs. never RRT) -Escalated | 0.80 [0.78 – 0.81] | 0.38 | 0.83 | 0.15 | 0.94 |
| Any alert (vs. No alert) - Escalated | 0.45 [0.43 – 0.47] | 0.63 | 0.44 | 0.08 | 0.94 |
| RRT only (vs. No Alert) - Escalated | 0.75 [0.73 – 0.77] | 0.35 | 0.78 | 0.11 | 0.94 |
| Primary only (vs. No Alert) - Escalated | 0.52 [0.50 – 0.54] | 0.40 | 0.53 | 0.05 | 0.94 |
| Both Alerts only (vs. No Alert) - Escalated | 0.85 [0.83 – 0.87] | 0.32 | 0.90 | 0.22 | 0.94 |
| Ever RRT (vs. never RRT) - ICU | 0.81 [0.79 – 0.82] | 0.43 | 0.83 | 0.11 | 0.97 |
| Any alert (vs. No alert) - ICU | 0.45 [0.43 – 0.47] | 0.66 | 0.44 | 0.06 | 0.96 |
| RRT only (vs. No Alert) - ICU | 0.76 [0.74 – 0.79] | 0.40 | 0.78 | 0.09 | 0.96 |
| Primary only (vs. No Alert) - ICU | 0.53 [0.51 – 0.55] | 0.40 | 0.53 | 0.03 | 0.96 |
| Both Alerts only (vs. No Alert) - ICU | 0.87 [0.85 – 0.88] | 0.38 | 0.89 | 0.17 | 0.96 |
| **Alert Level** | | | | | |
| RRT alert (vs. Primary and No alert) - Escalated | 0.78 [0.76 – 0.79] | 0.36 | 0.82 | 0.15 | 0.93 |
| RRT alert (vs. No alert) - Escalated | 0.68 [0.66 – 0.70] | 0.44 | 0.69 | 0.15 | 0.94 |
| Any alert (vs. No alert) - Escalated | 0.43 [0.41 – 0.45] | 0.70 | 0.41 | 0.10 | 0.94 |
| RRT alert (vs. Primary and No alert) - ICU | 0.79 [0.78 – 0.81] | 0.40 | 0.82 | 0.12 | 0.96 |
| RRT alert (vs. No alert) - ICU | 0.68 [0.66 - 0.70] | 0.61 | 0.69 | 0.12 | 0.96 |
| Any alert (vs. No alert) - ICU | 0.43 [0.41 – 0.45] | 0.74 | 0.41 | 0.07 | 0.96 |

*PPV: Positive Predictive Value = True Positive/ (True Positive + False Positive)

**NPV: Negative Predictive Value = True Negative/ (False Negative + True Negative)
